# Longitudinal Trends in Secondary Cardiovascular Prevention After Peripheral Artery Disease Diagnosis

**DOI:** 10.64898/2026.11.19.26368543

**Authors:** Karoline Kallis, Pooja Kumar, Eastern Kang-Sim, Nicholas Neel, John Z. Deng, Chyler Quitevis, Tom Alsaigh, Elsie G. Ross

**Affiliations:** Department of Surgery, Division of Surgical Sciences, University of California San Diego School of Medicine, La Jolla, CA, USA; Department of Surgery, Division of Vascular Surgery, University of California, San Diego School of Medicine, La Jolla, CA, USA

**Keywords:** peripheral artery disease, electronic health record, Epic Cosmos, guideline-directed medication therapy, real world data

## Abstract

**Background:** Peripheral artery disease (PAD) is associated with substantial cardiovascular and limb-related morbidity, making long-term cardiovascular prevention an important part of care. However, longitudinal patterns of guideline-based quality measure achievement after PAD diagnosis remain poorly characterized in broad clinical populations.

**Methods:** We conducted a retrospective cohort study using Epic Cosmos electronic health record (EHR) data to identify adults aged 18 to 75 years with PAD diagnosed from 2019 to 2023. Four cardiovascular prevention components based on Merit-based Incentive Payment System (MIPS) Quality Measures were assessed annually after PAD diagnosis: blood pressure target achievement, documented tobacco-free status, antiplatelet therapy, and statin therapy. Complete achievement was defined as meeting all four measures. We assessed quality-measure achievement across groups, three-year quality-index trends, and associations between complete achievement and subsequent outcomes.

**Results:** Among 906,686 patients, model-adjusted first-year achievement was 60.6% for tobacco-free status, 61.9% for blood pressure control, 39.5% for antiplatelet therapy, and 63.4% for statin therapy; 10.4% achieved all four. Model-adjusted complete achievement was 12.4% in chronic limb-threatening ischemia (CLTI), 10.5% in claudication, and 10.2% in unspecified PAD. Model-adjusted rates varied by sex, race/ethnicity, insurance type, rurality, PAD severity, and comorbidities and ranged from 7.6% to 14.7%. Complete achievement in year 1 was associated with lower adjusted odds of amputation (odds ratio (OR), 0.91), vascular procedures (OR, 0.86), stroke (OR, 0.90), and myocardial infarction (OR, 0.94).

**Conclusions:** In this large national EHR cohort, individual quality measures improved after PAD diagnosis, but only 10.4% of patients achieved all four assessed components during the first year after diagnosis. Differences across quality domains and patient subgroups highlight variation in longitudinal cardiovascular risk prevention, particularly among patients with less clinically apparent disease.

## Introduction

Peripheral artery disease (PAD) affects an estimated 8.5 million adults in the United States and is associated with increased risk of major adverse cardiovascular events, major adverse limb events, functional decline, and impaired quality of life.^1–6^ Although guideline-recommended therapies are associated with improved cardiovascular and limb-related outcomes, implementation in routine care remains incomplete. Prior studies have shown that PAD patients are less likely to receive optimal medical therapy than patients with coronary artery disease (CAD), despite both conditions representing clinical atherosclerotic cardiovascular disease. These findings highlight persistent gaps in cardiovascular prevention after PAD diagnosis.^2,7–10^

National quality programs like Merit-based Incentive Payment System (MIPS) have defined performance measures to promote standardized medical management among patients with ischemic vascular disease including PAD. MIPS Quality Measure #441 defines optimal control as achievement of four targets: blood pressure ≤140/90 mmHg, documented tobacco-free status, daily aspirin or another antiplatelet agent unless contraindicated, and statin therapy unless contraindicated. Together, these components provide a standardized framework for assessing cardiovascular risk-factor management after PAD diagnosis.^11^

Furthermore, PAD is a heterogeneous disease with variable clinical presentation and severity ranging from asymptomatic disease to claudication and chronic limb-threatening ischemia (CLTI). Prior studies suggest that care is not delivered uniformly across patient phenotypes and differences in diagnosis, treatment, and medication use have been reported by sex, race and ethnicity, socioeconomic status, insurance coverage, and clinical presentation.^12,13^

Additionally, longitudinal evidence on guideline-concordant care after PAD diagnosis remains limited. Most studies have examined selected populations, specific care settings, or a single time point. Therefore, less is known about the factors associated with individual measure achievement and whether differences in care persist over time.^9,10,13,14^

In this study, we evaluated achievement of selected secondary cardiovascular prevention measures among patients with PAD in a large national EHR cohort. Using MIPS Quality Measure #441 as a framework, we assessed its four components during the first year after PAD diagnosis: blood pressure target achievement, documented tobacco-free status, antiplatelet therapy, and statin therapy. We also examined demographic, socioeconomic, and clinical factors associated with component achievement, longitudinal changes over three years, and associations between complete achievement and subsequent clinical outcomes.

## Materials and Methods

### Study Population

This retrospective cohort study used EHR data from Epic Cosmos, a nationally representative database including de-identified data from more than 300 million patients across more than 1,800 hospitals in the United States. The study cohort included patients with at least two PAD diagnosis codes recorded between 2019 and 2023, occurring within a two-year period and separated by at least 30 days. The complete list of diagnosis codes used to identify PAD is provided in **Supplemental Table S1**. The index date was defined as the date of the second qualifying PAD diagnosis code. Eligible patients were required to be 18 to 75 years of age at both qualifying PAD diagnosis codes, receive care in the United States, and remain active in the registry for at least three years after the index date. They also needed documentation of blood pressure or medication therapy in the year prior index date and in at least one follow-up year. Active status was defined as having at least two encounters within a two-year period. Patients with missing data on sex, race/ethnicity, insurance category, SVI, or RUCA classification were excluded, as were those receiving care through Veterans Affairs healthcare systems.

### Assessment of Quality Measures

At the index date, baseline achievement of each quality measure was determined using information documented during the preceding 12 months. Following index date, quality measures were assessed annually during the first three years. Measures were selected based on MIPS Quality Measure #441 and included blood pressure target achievement, documented tobacco-free status, antiplatelet therapy, and statin therapy.^11^ Achievement of the blood pressure target was defined as a mean blood pressure of ≤140/90 mmHg during the corresponding follow-up year. Tobacco-free status was defined using documented smoking status during each annual measurement period. Patients with a documented status of former smoker or never smoker were classified as meeting the tobacco-free quality measure, whereas current smokers and patients without a documented smoking status were classified as not meeting the measure. Because smoking status was not documented at every encounter, missing smoking status in earlier measurement periods was backfilled using later documented smoking status. Missing smoking status in earlier measurement periods was backfilled using later documentation of never-smoking or current-smoking status, provided no conflicting earlier status was recorded. Patients were considered to be receiving antiplatelet (including aspirin) or statin therapy during a given follow-up year if a qualifying prescription was ordered during that year or a previously ordered prescription remained active during that year according to the recorded end date. Only records classified as prescribed medications were included in the analysis and antiplatelet prescriptions categorized under non-cardiovascular related therapeutic classes were excluded.

Quality measure achievement was evaluated separately for each measure and collectively as a composite quality index (QI, 0–100%), representing the proportion of the four MIPS-informed care targets achieved by each patient. Achievement rates were calculated at baseline and annually during the first three years following PAD diagnosis. Longitudinal trends in blood pressure control, tobacco-free status, antiplatelet therapy, and statin therapy were evaluated descriptively and stratified by PAD severity (claudication, CLTI, and unspecified). Among patients without CLTI at baseline, we identified subsequent documentation of claudication or CLTI during follow-up using diagnosis codes recorded after the index date. We separately assessed documented smoking cessation counseling after the index date among patients who did not meet the tobacco-free measure at baseline.

### Factors Associated with First-Year Quality Measure Achievement

The first-year analyses included 904,898 patients with at least one quality measure recorded during the first year after PAD diagnosis. Multivariable logistic regression models were used to identify demographic, socioeconomic, and clinical factors associated with achievement of each quality measure. Separate models were fit for blood pressure control, tobacco-free status, antiplatelet therapy, statin therapy, and complete achievement of all four measures (QI = 100%). We used the fitted models to estimate achievement rates for each subgroup after adjusting for the other patient characteristics included in the models.

Covariates included age group, sex, race/ethnicity, insurance type, SVI quartile, RUCA classification, PAD severity, diabetes, hypertension, hyperlipidemia, end-stage renal disease, obesity, chronic obstructive pulmonary disease, coronary artery disease, cerebrovascular disease, and heart failure. Age at index was categorized as <50, 50–64, and 65–75 years to facilitate interpretation of subgroup differences. Unadjusted first-year achievement rates are reported in **Supplemental Table S7**, model-adjusted rates in **Table 2**, and adjusted odds ratios (ORs) with 95% confidence intervals (CIs) in **Supplemental Table S8**.

### Longitudinal Changes in Quality Measure Achievement

A linear mixed-effects model was used to evaluate longitudinal changes in the composite quality index (QI) during the first three years following PAD diagnosis. The QI was modeled as a function of follow-up time, race/ethnicity, insurance type, sex, and PAD severity, with interaction terms between follow-up time and each subgroup variable to assess differences in quality measure trajectories over time.

The model was adjusted for age, SVI, RUCA classification, diabetes, hypertension, hyperlipidemia, end-stage renal disease, obesity, chronic obstructive pulmonary disease, coronary artery disease, cerebrovascular disease, and heart failure. Patient-specific random intercepts were included to account for repeated measurements within individuals. Adjusted marginal means and 95% CI were estimated from the fitted model and used to visualize longitudinal trends across demographic and clinical subgroups.

### Association Between Quality Measure Achievement and Clinical Outcomes

Multivariable logistic regression models were used to evaluate associations between achievement of all four QI measures during the assessment period and subsequent PAD-related and cardiovascular events. Outcomes included lower extremity amputation (**Supplemental Table S4),** vascular procedures (**Supplemental Table S4**), stroke (**Supplemental Table S6**), and acute myocardial infarction (AMI, **Supplemental Table S5**). For each clinical outcome, we identified the first recorded event after the index date. Separate models were fitted for each outcome using two QI assessment periods after diagnosis: year 1 alone and years 1–2 combined. We used landmark analyses to limit immortal time bias. The 1-year analysis included patients who remained alive and free of the outcome through year 1 and had quality-measure data available for that year. The 2-year analysis required survival without the outcome through year 2 and quality-measure data in both years. Complete achievement required meeting all four measures in each year of the corresponding assessment period. Models were adjusted for age, sex, race/ethnicity, insurance type, SVI quartile, PAD severity, diabetes, hypertension, hyperlipidemia, end-stage renal disease, obesity, chronic obstructive pulmonary disease, coronary artery disease, cerebrovascular disease, heart failure, and index year. Associations are reported as ORs with 95% CIs.

Cohort identification was performed using SQL queries. Statistical analyses and visualizations were conducted using custom scripts implemented in Python (version 3.13.9).

## Results

The final cohort comprised 906,686 patients with PAD (**Figure 1**). Quality-metric documentation was available at baseline and through year 1 for 904,898 patients (99.8% of the cohort), through year 2 for 902,458 (99.5%), and through year 3 for 892,045 (98.4%). Mean age at index was 64.0 years; 55.8% were male and 70.7% were White. At baseline, PAD severity was classified as claudication in 8.9% of patients, CLTI in 7.9%, and unspecified PAD in 83.2% (**Table 1**). During follow-up, 16.7% of patients with claudication at baseline subsequently had documented CLTI. Among patients with unspecified PAD at baseline, 6.5% subsequently had documented claudication and 8.1% had documented CLTI.

**Figure 1.**
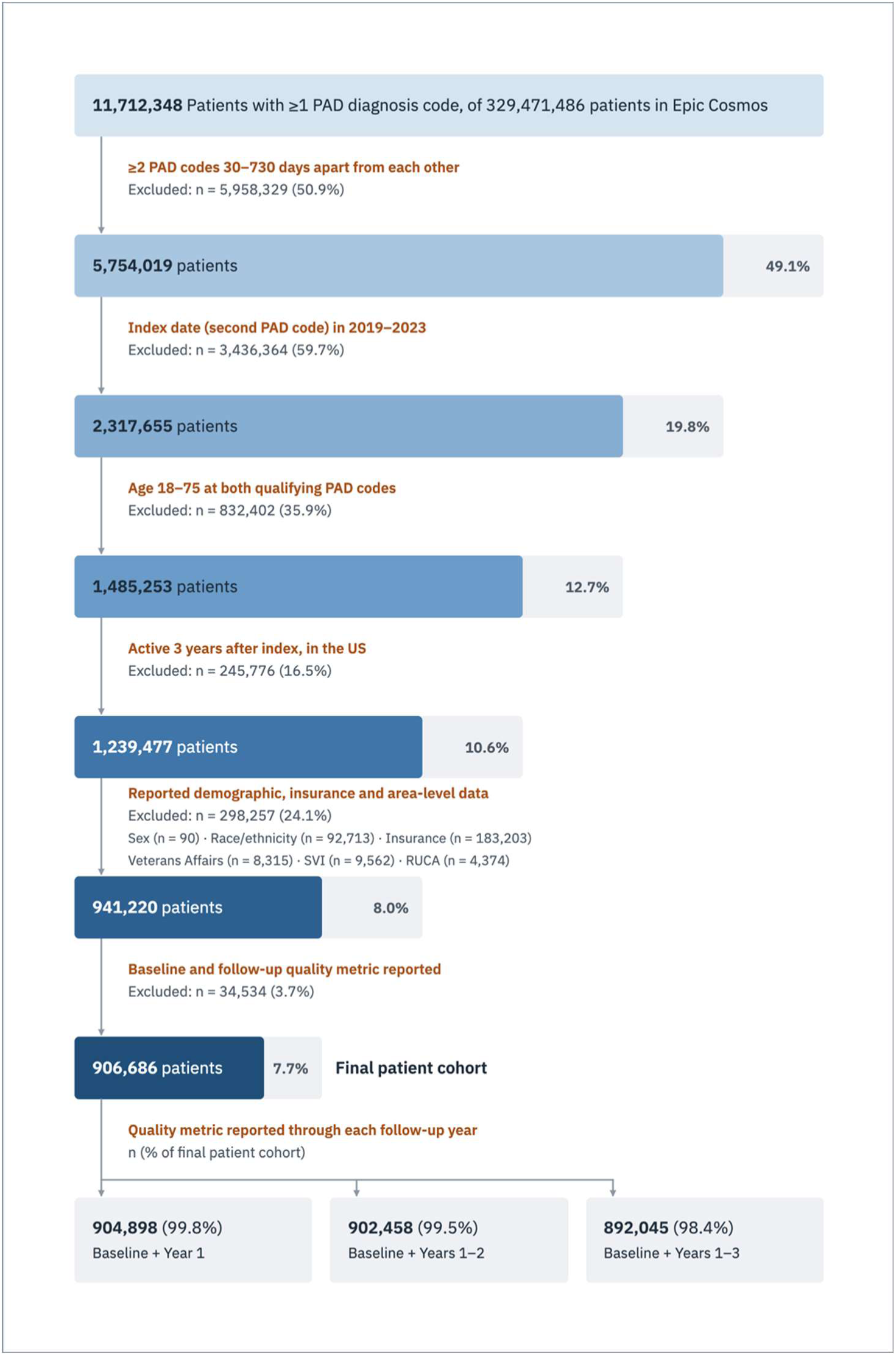
Patient selection flow diagram. Baseline was defined as the year before the index date, which was the date of the second qualifying PAD code. Percentages for patients retained at each selection step are relative to the initial 11,712,348 patients with at least one PAD code. Percentages for patients excluded are relative to the number entering that selection step. The lower boxes show availability of quality-metric documentation through follow-up years 1–3, with percentages relative to the final study cohort. PAD, peripheral artery disease; SVI, Social Vulnerability Index; RUCA, Rural-Urban Commuting Area.

**Table 1.** Baseline patient characteristics. PAD, peripheral artery disease; MA, Medicare Advantage; MR, Medicaid Replacement; SVI, Social Vulnerability Index; RUCA, Rural-Urban Commuting Area; DM, diabetes mellitus; CAD, coronary artery disease; HF, heart failure; HTN, hypertension; HLD, hyperlipidemia; ESRD, end-stage renal disease; COPD, chronic obstructive pulmonary disease; CVA, cerebrovascular accident; CLTI, chronic limb-threatening ischemia. Self-pay/uninsured patients are reported separately from Other insurance.

| **Parameter** | **Statistic** | **Total** |
|---|---|---|
| Number of patients | n | 906,686 |
| Age at index | mean ± SD | 64.0 ± 9.0 y |
| Race/Ethnicity | n (%) Black | 158,720 (17.5%) |
|  | n (%) White | 641,431 (70.7%) |
|  | n (%) Asian | 21,158 (2.3%) |
|  | n (%) Hispanic | 65,651 (7.2%) |
|  | n (%) Other | 19,726 (2.2%) |
| Sex | n (%) Female | 400,687 (44.2%) |
|  | n (%) Male | 505,999 (55.8%) |
| Insurance type | n (%) MA | 248,679 (27.4%) |
|  | n (%) MR | 73,979 (8.2%) |
|  | n (%) Commercial | 223,293 (24.6%) |
|  | n (%) Medicare | 293,755 (32.4%) |
|  | n (%) Medicaid | 38,080 (4.2%) |
|  | n (%) Self-pay/uninsured | 15,499 (1.7%) |
|  | n (%) Other | 13,401 (1.5%) |
| SVI | n (%) Q1 | 128,263 (14.1%) |
|  | n (%) Q2 | 183,182 (20.2%) |
|  | n (%) Q3 | 233,521 (25.8%) |
|  | n (%) Q4 | 361,720 (39.9%) |
| RUCA classification | n (%) Metropolitan | 734,895 (81.1%) |
|  | n (%) Micropolitan | 94,064 (10.4%) |
|  | n (%) Small town | 48,084 (5.3%) |
|  | n (%) Rural | 29,643 (3.3%) |
| Elixhauser comorbidity score | n (%) 0 | 136,919 (15.1%) |
|  | n (%) 1-2 | 308,411 (34.0%) |
|  | n (%) 3-5 | 115,817 (12.8%) |
|  | n (%) 6+ | 345,539 (38.1%) |
| Comorbidity | n (%) DM | 443,030 (48.9%) |
|  | n (%) CAD | 353,457 (39.0%) |
|  | n (%) HF | 190,977 (21.1%) |
|  | n (%) HTN | 719,933 (79.4%) |
|  | n (%) HLD | 657,368 (72.5%) |
|  | n (%) ESRD | 48,454 (5.3%) |
|  | n (%) Obesity | 313,462 (34.6%) |
|  | n (%) COPD | 159,761 (17.6%) |
|  | n (%) CVA | 159,789 (17.6%) |
| PAD severity | n (%) CLTI | 71,645 (7.9%) |
|  | n (%) Claudication | 80,916 (8.9%) |
|  | n (%) Unspecified | 754,125 (83.2%) |

**Table 2.** Adjusted first-year PAD quality measure achievement. PAD, peripheral artery disease; BP, blood pressure; QI, quality index (QI = 100% indicates achievement of all four measures); MA, Medicare Advantage; MR, Medicaid Replacement; SVI, Social Vulnerability Index; Q1–Q4, quartiles 1–4; RUCA, Rural-Urban Commuting Area; DM, diabetes mellitus; CAD, coronary artery disease; HF, heart failure; HTN, hypertension; HLD, hyperlipidemia; ESRD, end-stage renal disease; COPD, chronic obstructive pulmonary disease; CVA, cerebrovascular accident; CLTI, chronic limb-threatening ischemia.

| **Parameter** | **Statistic** | **Tobacco-free status** | **BP control** | **Antiplatelet** | **Statin** | **QI = 100%** |
|---|---|---|---|---|---|---|
| Proportion of patients | % | 60.6% | 61.9% | 39.5% | 63.4% | 10.4% |
| Age group | <50 | 56.5% | 66.4% | 37.1% | 55.3% | 8.4% |
|  | 50–64 | 54.4% | 62.3% | 41.7% | 63.9% | 9.9% |
|  | 65–75 | 65.0% | 61.1% | 38.4% | 64.2% | 11.0% |
| Race/Ethnicity | Black | 59.9% | 54.7% | 41.8% | 65.0% | 9.8% |
|  | White | 59.2% | 63.5% | 38.5% | 62.7% | 9.9% |
|  | Asian | 73.8% | 64.8% | 40.3% | 66.7% | 14.7% |
|  | Hispanic | 71.5% | 62.6% | 42.7% | 66.2% | 14.7% |
|  | Other | 63.0% | 62.9% | 40.5% | 61.3% | 11.5% |
| Sex | Female | 62.1% | 62.5% | 37.6% | 61.8% | 9.8% |
|  | Male | 59.4% | 61.4% | 40.9% | 64.7% | 10.9% |
| Insurance type | MA | 59.2% | 64.4% | 38.1% | 62.8% | 10.2% |
|  | MR | 52.0% | 63.1% | 46.3% | 65.4% | 11.2% |
|  | Commercial | 64.3% | 61.8% | 39.1% | 65.7% | 11.3% |
|  | Medicare | 62.3% | 60.0% | 37.7% | 60.9% | 9.6% |
|  | Medicaid | 52.9% | 61.1% | 46.4% | 66.4% | 11.4% |
|  | Self-pay/uninsured | 56.4% | 56.0% | 48.4% | 73.0% | 12.2% |
|  | Other | 61.5% | 61.8% | 40.0% | 62.8% | 10.8% |
| SVI | Q1 | 63.4% | 62.9% | 37.7% | 64.8% | 10.6% |
|  | Q2 | 61.9% | 62.5% | 38.2% | 63.3% | 10.3% |
|  | Q3 | 60.6% | 62.1% | 39.4% | 63.2% | 10.4% |
|  | Q4 | 58.9% | 61.2% | 40.7% | 63.2% | 10.5% |
| RUCA classification | Metropolitan | 61.2% | 62.1% | 39.5% | 64.0% | 10.6% |
|  | Micropolitan | 58.7% | 61.5% | 39.6% | 61.9% | 9.8% |
|  | Small town | 58.0% | 60.5% | 38.7% | 60.5% | 9.0% |
|  | Rural | 57.0% | 60.7% | 40.0% | 59.7% | 9.1% |
| Comorbidity | DM | 63.3% | 60.4% | 39.8% | 67.1% | 11.2% |
|  | CAD | 60.5% | 62.6% | 50.1% | 66.5% | 14.2% |
|  | HF | 64.9% | 65.0% | 41.9% | 63.0% | 12.2% |
|  | HTN | 60.7% | 60.3% | 40.0% | 64.5% | 10.5% |
|  | HLD | 61.0% | 63.1% | 39.6% | 70.4% | 11.2% |
|  | ESRD | 74.0% | 52.3% | 40.7% | 53.3% | 10.2% |
|  | Obesity | 67.1% | 65.1% | 36.1% | 63.4% | 11.3% |
|  | COPD | 42.5% | 64.2% | 41.2% | 63.3% | 7.6% |
|  | CVA | 59.9% | 60.1% | 49.4% | 67.3% | 13.2% |
| PAD severity | CLTI | 55.0% | 60.3% | 57.8% | 60.6% | 12.4% |
|  | Claudication | 52.6% | 57.8% | 52.1% | 61.9% | 10.5% |
|  | Unspecified | 62.0% | 62.5% | 36.3% | 63.9% | 10.2% |

### First-Year Quality Measure Achievement

During the first year after PAD diagnosis, model-adjusted achievement was highest for statin therapy (63.4%), followed by blood pressure control (61.9%) and documented tobacco-free status (60.6%). Adjusted antiplatelet achievement was 39.5%, and the adjusted estimate for complete achievement of all four measures was 10.4% (**Table 2**).

Model-adjusted complete achievement remained low across subgroups (**Table 2**). It ranged from 9.8% in Black patients to 14.7% in Asian and Hispanic patients and from 10.2% in unspecified PAD to 12.4% in CLTI. Among patients with comorbidities, model-adjusted complete achievement ranged from 7.6% in those with chronic obstructive pulmonary disease to 14.2% in those with coronary artery disease. For individual measures, blood pressure control was 54.7% in Black patients versus 63.5% in White patients. Antiplatelet achievement was 36.3% in unspecified PAD versus 52.1% in claudication and 57.8% in CLTI.

### Longitudinal Trends in Individual Quality Measure Achievement

Documented blood pressure control rose slightly from baseline to year 1 and then declined through year 3 across PAD severity groups (**Figure 2**). From baseline to year 3, achievement decreased from 56.2% to 53.5% among patients with claudication, from 58.2% to 49.4% among those with CLTI, and from 61.7% to 58.4% among those with unspecified PAD.

**Figure 2.**
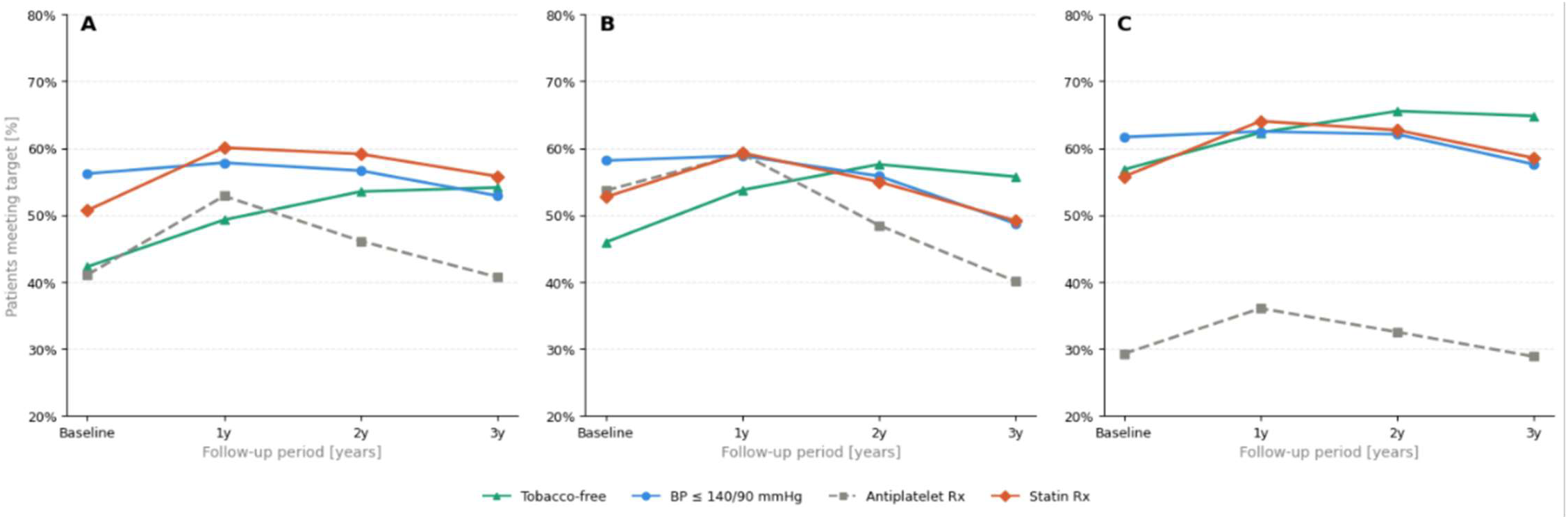
Longitudinal achievement of individual quality measures by peripheral artery disease (PAD) severity. Panels show quality measure achievement A) claudication, B) chronic limb-threatening ischemia (CLTI), and C) unspecified PAD at baseline and in years 1–3 after the index diagnosis. Baseline is defined as the preceding 12 months prior index date. Tobacco-free denotes documented tobacco-free status; BP, blood pressure; Rx, prescription.

Documented tobacco-free status increased from baseline to year 3 in all groups: from 42.3% to 54.7% among patients with claudication, from 45.9% to 56.5% among those with CLTI, and from 56.8% to 65.7% among those with unspecified PAD. Among patients who did not meet the documented tobacco-free measure at baseline, 12.9% had smoking cessation counseling recorded within three years after the index date.

Documented antiplatelet achievement was lowest among patients with unspecified PAD at every time point. From year 1 to year 3, achievement decreased from 53.0% to 41.2% in claudication, from 59.2% to 40.6% in CLTI, and from 36.1% to 29.3% in unspecified PAD.

Patients with CLTI had the highest year-1 rate, whereas patients with claudication had a slightly higher year-3 rate.

Documented statin achievement increased from baseline to year 1 and then decreased through year 3 in all three groups. Year-3 rates were 56.4% for claudication, 49.8% for CLTI, and 59.3% for unspecified PAD, compared with year-1 rates of 60.2%, 59.4%, and 64.2%, respectively.

### Longitudinal Patterns in Composite Quality Measure Achievement

Adjusted composite QI improved after PAD diagnosis across all subgroups, with relatively modest differences between patient groups (**Figure 3**). QI increased from baseline to year 1 in every racial and ethnic group and declined by year 3, although all year-3 estimates remained above baseline (**Figure 3A**). Asian patients had the highest adjusted QI at each time point (57.5% at baseline, 64.1% in year 1, and 59.9% in year 3), whereas Black patients had the lowest (50.6%, 58.3%, and 54.6%, respectively). Hispanic patients had the second-highest year-3 estimate (58.9%); estimates were 55.4% among White patients and 54.9% among patients in the Other group.

**Figure 3.**
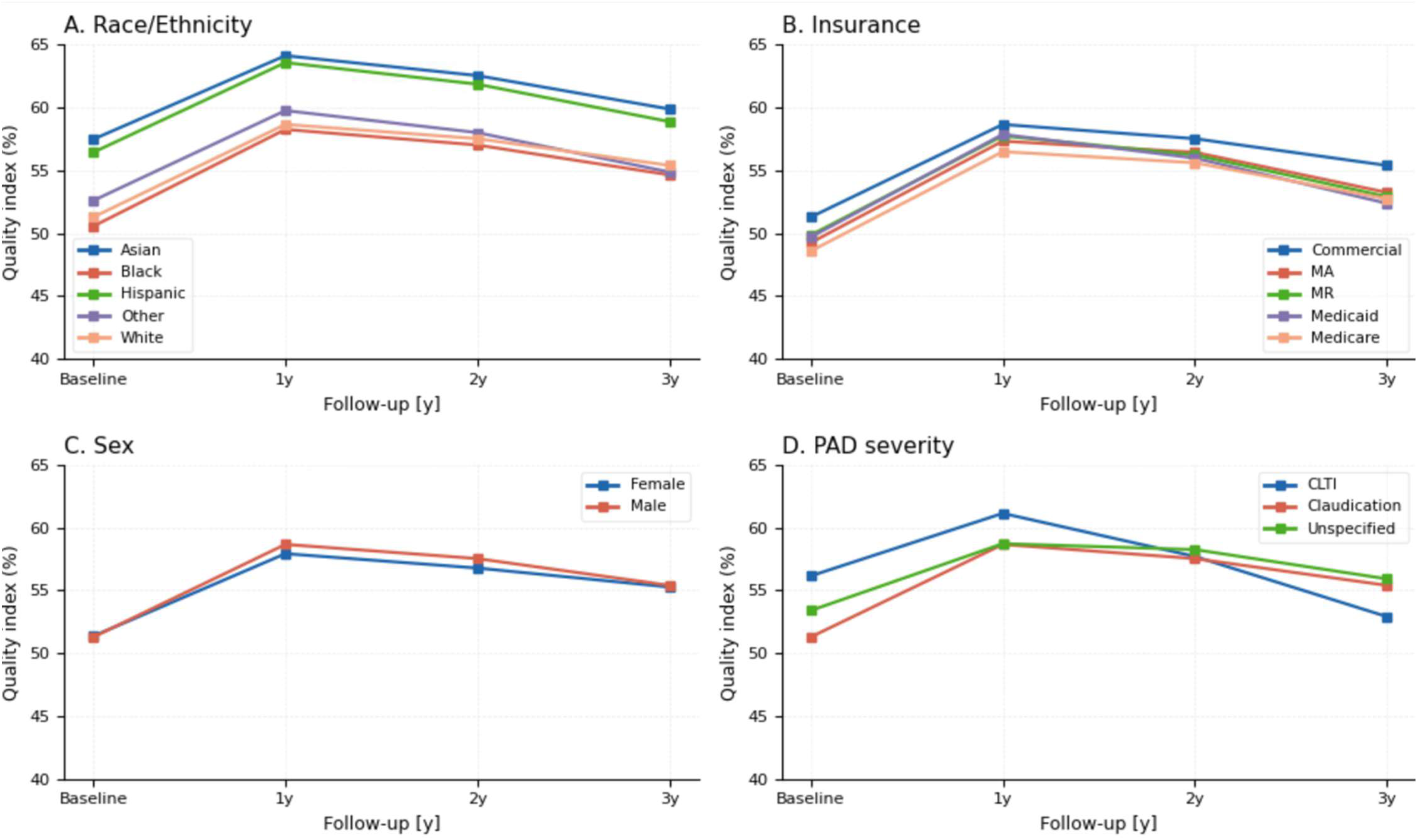
Adjusted composite quality index trajectories by patient subgroup. QI is the percentage of four prevention targets achieved; baseline is defined as the 12 months before the index PAD diagnosis. Panels show A) race/ethnicity, B) five insurance categories, C) sex, and D) PAD severity through year 3. Estimates are from a linear mixed-effects model with follow-up time, subgroup terms, and their interactions, adjusted for demographic, socioeconomic, geographic, and clinical covariates. PAD, peripheral artery disease; MA, Medicare Advantage; MR, Medicaid Replacement; CLTI, chronic limb-threatening ischemia.

Among the insurance groups shown in **Figure 3B**, adjusted QI rose from baseline to year 1 and then declined through year 3. Commercially insured patients had the highest estimates at every time point, increasing from 51.3% at baseline to 58.7% in year 1 and then declining to 55.4% in year 3. Medicare patients had the lowest estimate at baseline (48.6%) and in years 1 and 2 (56.5% and 55.6%), whereas Medicaid patients had the lowest year-3 estimate (52.4%).

Adjusted QI was similar by sex at baseline (51.4% for women and 51.3% for men; **Figure 3C**). It increased to 57.9% and 58.7%, respectively, in year 1, then declined to 55.3% and 55.4% by year 3.

Patients with CLTI had the highest adjusted QI at baseline (56.1%) and year 1 (61.1%; **Figure 3D**), followed by a decline to 52.9% in year 3. Patients with claudication increased from 51.3% at baseline to 58.7% in year 1 and ended at 55.4% in year 3. Patients with unspecified PAD increased from 53.4% at baseline to 58.7% in year 1 and had the highest year-2 and year-3 estimates (58.2% and 55.9%).

### Association of Complete Quality Measure Achievement With PAD-related Outcomes

Clinical outcome rates were highest among patients with CLTI for all five outcomes. Major and minor amputation rates were 17.5% and 15.8% in CLTI, compared with 3.3% and 2.8% in claudication and 3.4% and 3.3% in unspecified PAD. Vascular procedure rates were 30.9%, 23.9%, and 7.2% across CLTI, claudication and unspecified PAD, respectively. Myocardial infarction rates were 24.3%, 19.7%, and 17.3%, and stroke rates were 18.4%, 14.7%, and 14.6%, respectively.

Complete achievement of all four quality measures was associated with lower odds of each clinical outcome in adjusted analyses (**Table 3**). In the 1-year analysis, the ORs were 0.91 (95% confidence interval (CI), 0.88–0.95) for amputation, 0.86 (0.83–0.89) for vascular procedures, 0.90 (0.88–0.93) for stroke, and 0.94 (0.92–0.96) for acute myocardial infarction. When complete achievement was required during both years 1 and 2, the corresponding ORs were 0.84, 0.79, 0.86, and 0.90 for amputation, vascular procedures, stroke and myocardial infarction, respectively. Full adjusted model results are presented in **Supplemental Table S9**.

**Table 3.**
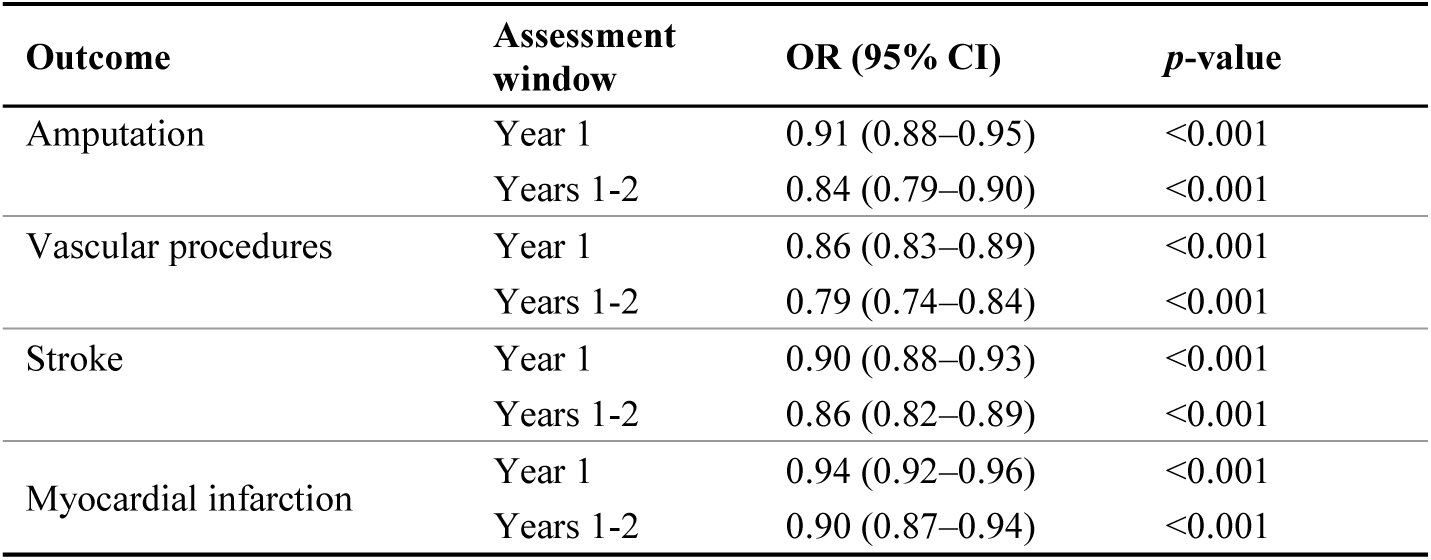
Association between complete quality index achievement and subsequent clinical outcomes. Odds ratios compare QI = 100% with QI <100% during year 1 or during both years 1 and 2; outcomes were assessed after each QI window. Models were adjusted for the covariates listed in Methods. CI, confidence interval; OR, odds ratio; QI, quality index.

## Discussion

In this national EHR cohort of 906,686 patients with PAD, we evaluated four components of cardiovascular prevention based on MIPS Quality Measure #441 during the first three years after diagnosis: blood pressure target achievement, documented tobacco-free status, antiplatelet therapy, and statin therapy. Overall, achievement of these four measures remained suboptimal throughout follow-up. Adjusted first-year achievement of individual measures ranged from 39.5% to 63.4%, whereas only 10.4% achieved all four MIPS quality targets during the first year after diagnosis. Quality improved after diagnosis, particularly for medication-based measures, but remained below 65% across all subgroups during follow-up. Meeting all four measures was associated with lower adjusted odds of amputation, vascular procedures, stroke, and AMI.

The low rate of complete quality measure achievement in our study is consistent with prior evidence that guideline-recommended PAD care remains underutilized. In the PORTRAIT registry of symptomatic patients receiving specialty vascular care, 89% received antiplatelet therapy and 83% received statin therapy, but only 19.7% achieved all four measures.^14^ Similarly, fewer than half of patients undergoing peripheral vascular intervention in the Vascular Quality Initiative registry received guideline-directed medical therapy.^9^ Although direct comparisons are limited by differences in cohort design, care setting, and quality measure definitions, our findings extend this prior work by showing that complete quality measure achievement remains low outside specialty and procedural cohorts.

Quality measure achievement was generally higher among patients with more advanced PAD and established cardiovascular comorbidities, suggesting that guideline-recommended therapies may be more consistently implemented when cardiovascular risk is more clinically apparent. Patients with CLTI had higher complete quality measure achievement than patients with claudication (12.4% vs. 10.5%) and had 28% higher odds of documented antiplatelet therapy. In contrast, patients with unspecified PAD had approximately half the odds of documented antiplatelet therapy compared with patients with claudication. Similar patterns were observed among patients with established cardiovascular comorbidities, consistent with prior national ambulatory care data showing that patients with both PAD and CAD were substantially more likely to receive antiplatelet therapy, statins, and smoking cessation counseling than patients with PAD alone.^15^ In our study only 12.9% of patients who did not meet the tobacco-free measure at baseline had smoking-cessation counseling documented within three years, suggesting a potential missed opportunity to support cessation, although incomplete documentation and the inclusion of patients with unknown smoking status limit interpretation. The lower achievement observed among patients with unspecified PAD was driven primarily by lower recorded antiplatelet prescription, which may partly reflect incomplete capture of over-the-counter medication. Achievement was also lower among patients with claudication than among those with CLTI, and 16.7% of patients with claudication at baseline subsequently had documented CLTI during follow-up. Among patients with unspecified PAD, 6.5% later had documented claudication and 8.1% later had documented CLTI. These findings suggest that unspecified PAD at diagnosis may reflect incomplete severity capture, delayed recognition, or disease progression, rather than uniformly less severe disease. Together, the lower achievement and subsequent documentation of more severe disease highlight potential missed opportunities for comprehensive cardiovascular risk reduction among patients with claudication or unspecified PAD.

Differences in individual measures suggest that gaps in cardiovascular prevention vary across patient groups. Women had lower documented antiplatelet and statin therapy, while adjusted blood pressure target achievement was lower among Black than White patients (54.7% versus 63.5%). Patients with Medicaid coverage had lower documented tobacco-free status despite higher antiplatelet achievement. These patterns are consistent with prior reports of sex, racial, socioeconomic, and geographic differences in PAD care.^12,13,16,17^ The subgroup differences we observed may reflect multiple factors, including delayed diagnosis, variation in symptom presentation, access to specialty care, medication cost, and other barriers to longitudinal risk-factor management.^12,13^ The measure-specific findings suggest that composite scores can obscure gaps in specific components of care, supporting quality improvement efforts tailored to the measures and populations with the largest differences.

Adjusted composite QI increased from baseline to year 1 and declined by year 3, although most year-3 estimates remained above baseline. Declines in blood pressure target achievement and documented antiplatelet and statin therapy may reflect challenges in sustaining preventive care over time, including treatment discontinuation, inconsistent follow-up, and incomplete capture of ongoing care. Aspirin use may be particularly under captured because nonprescription use may not be consistently documented in the EHR.

Meeting all four measures in year 1 was associated with 6% to 14% lower adjusted odds across the four considered outcomes, with the strongest inverse association for vascular procedures (OR, 0.86) and the weakest for AMI (OR, 0.94). Requiring complete achievement in both years 1 and 2 yielded ORs from 0.79 for vascular procedures to 0.90 for AMI. This pattern is consistent with the potential importance of sustained preventive care, although the observational design precludes causal interpretation. Although complete achievement of the four prevention measures was associated with lower odds of amputation, vascular procedures, stroke, and AMI, the composite measure primarily reflects cardiovascular prevention and does not capture several important components of limb-preservation care or the clinical indications for vascular procedures. These include foot surveillance, wound management, glycemic control, vascular evaluation, and revascularization when clinically indicated. Meeting the four cardiovascular prevention measures therefore complements, rather than replaces, targeted limb-preservation strategies for patients at high risk of amputation, particularly those with diabetes, kidney disease, or CLTI. ^18,19^

Strengths of this study include the large national EHR cohort of more than 900,000 patients with PAD and the inclusion of diverse health systems, clinical settings, payer types, racial and ethnic groups, and geographic regions. The longitudinal design allowed evaluation of changes in care over three years after diagnosis, while simultaneous assessment of four prevention measures enabled examination of both individual measures and complete achievement. This broad real-world population extends prior research based primarily on specialty registries and procedural cohorts and enabled comparisons across clinical, demographic, socioeconomic, insurance, and geographic subgroups, which contributes to established PAD research.^20^

This study should be interpreted in the context of its limitations. First, quality measure achievement was based on EHR documentation and may not fully reflect care received or medications taken. Medication measures were based on prescriptions, and we could not determine whether patients filled prescriptions, took medications as prescribed, or discontinued therapy. Antiplatelet therapy may be particularly under captured because aspirin is often obtained over the counter and may not be consistently documented in the EHR. Smoking status is also imperfectly captured in EHR data, as it may be missing, inconsistently updated, or not reflect changes in tobacco use over time. Similarly, blood pressure control may be influenced by visit frequency, measurement conditions, acute illness, and the availability of recorded values. Further, although MIPS Quality Measure #441 captures important cardiovascular prevention measures, it does not encompass all aspects of contemporary PAD care, including intensive lipid management, low-dose rivaroxaban when appropriate, diabetes medications with cardiovascular benefit, and structured exercise therapy.

Further, PAD diagnosis and severity were based on diagnosis codes, which may not fully capture symptom burden, anatomic disease severity, wound status, ankle-brachial index results, or clinical decision-making. Patients classified as having unspecified PAD may include individuals with less clinically apparent disease or incomplete severity documentation, and subsequent changes in coded severity may reflect true progression or delays in clinical recognition and documentation.

Because inclusion required evidence of continued health system activity, the cohort may underrepresent patients with fragmented care, limited access, or loss to follow-up. This is particularly relevant because quality measure achievement may itself be influenced by healthcare engagement, encounter frequency, and documentation completeness. Requiring three years of active registry status may select patients with greater survival and continued healthcare engagement. Further, outcomes were identified only from recorded health-system events, with no censoring for loss to follow-up, variable follow-up duration by index date, and no accounting for the competing risk of death, which may have biased the observed associations.

Furthermore, we did not capture medication contraindications, intolerance, patient preferences, or clinical reasons for not pursuing specific treatment targets. As a result, complete quality measure achievement should not be interpreted as a realistic target of 100% across all patients, but rather as a population-level indicator of guideline-concordant cardiovascular prevention and potential gaps in care. Also, race and ethnicity were based on EHR-recorded categories, which generally reflect self-reported patient information but may also be affected by documentation practices and differences in how categories are collected across health systems. SVI and RUCA are area-level measures and may not capture individual socioeconomic circumstances, transportation barriers, medication affordability, or access to specialty care. The outcome analyses required patients to remain event-free during the quality-measure assessment period. The findings therefore apply only to patients who remained event-free through the end of that period and may not generalize to those who experienced earlier events. Residual differences in healthcare engagement, access to care, disease severity, and documentation may also have contributed to the observed associations.

Finally, because the study included patients diagnosed between 2019 and 2023, some observations overlapped with the COVID-19 pandemic and subsequent recovery period. Healthcare disruptions during this time may have affected visit frequency, blood pressure measurement, smoking status documentation, medication initiation, and timing of vascular evaluation or procedures, particularly among patients diagnosed early in the study period. However, the longitudinal design also captured care patterns beyond the initial pandemic period.

Overall, in this national EHR cohort of 906,686 patients with PAD, the model-adjusted estimate for complete achievement of all four prevention measures during the first year was 10.4%. Achievement differed by measure and subgroup. Compared with patients with claudication, those with unspecified PAD had lower adjusted odds of antiplatelet achievement but higher adjusted odds of statin achievement. These measure-specific findings support quality improvement efforts that address gaps in cardiovascular prevention across broad PAD populations. Meeting all four measures was associated with lower odds of amputation, vascular procedures, stroke, and AMI. These associations support further evaluation of the composite as a marker of cardiovascular prevention, alongside targeted limb-preservation strategies for patients at high risk of amputation.

## Data Availability

The patient-level electronic health record data used in this study were obtained from Epic Cosmos and are subject to Epic Cosmos data-use restrictions. The authors cannot share these data, and they are not publicly available.

## Acknowledgements

The authors would like to thank Ruth Laverde and Matthew Heard for their support with data access, technical infrastructure, and project coordination related to this study. The authors would like to acknowledge the use of ChatGPT (version GPT-6, OpenAI) for assistance with refining the language of the manuscript. The authors reviewed and edited the content and take full responsibility for the publication’s content.

